# Trends and Future Burden of Major Gastrointestinal Cancers in Jiangsu Province, China, 2010–2030

**DOI:** 10.64898/2026.07.16.26358207

**Authors:** Yanzheng Zou, Wei Wang, Luqiu Tao, Hongru Zhu, Haiyan Ju, Li Pan, Wei Wang

## Abstract

**Aim:** To assess temporal trends in incidence and mortality and project the future burden of five major gastrointestinal cancers in Jiangsu Province, China.

**Methods:** Population-based cancer registry data from Jiangsu Province between 2010 and 2021 were used to analyze the burden of esophageal, gastric, colon, rectal, and liver cancers. Age-standardized incidence and mortality rates were calculated and compared by cancer type, sex, and urban-rural residence. Joinpoint regression was used to estimate annual percentage changes (APC) and average annual percentage changes (AAPC). The APC from the most recent Joinpoint segment was used to project incidence and mortality rates to 2030.

**Results:** In 2021, gastric cancer had the highest age-standardized incidence and mortality among the five cancers. Incidence and mortality were consistently higher in males than in females and increased markedly after 50 years of age. From 2010 to 2021, age-standardized incidence and mortality declined for esophageal, gastric, and liver cancer, but increased for colon and rectal cancer. Colon cancer showed the steepest increase in both incidence and mortality. Rural areas experienced faster increases in colon and rectal cancer burden than urban areas. Projections to 2030 suggest continued declines in esophageal, gastric, and liver cancer, while colon cancer incidence and mortality are expected to rise further.

**Conclusion:** Jiangsu Province is experiencing a transition in gastrointestinal cancer burden, with continued declines in esophageal, gastric, and liver cancers but an emerging and growing burden of colorectal cancer, especially colon cancer. Prevention strategies should focus on expanding colorectal cancer screening and early diagnosis, particularly in rural areas, while sustaining control of esophageal, gastric, and liver cancers.

## 1. Introduction

Gastrointestinal cancers remain a major public health challenge in China. In 2022, colorectal cancer accounted for an estimated 517,100 new cases and 240,000 deaths, liver cancer for 367,700 new cases and 316,500 deaths, gastric cancer for 358,700 new cases and 260,400 deaths, and esophageal cancer for 224,000 new cases and 187,500 deaths^1^. Together, these cancers contribute substantially to the national cancer burden. Given their high incidence and mortality, continuous surveillance is needed to better understand their patterns and to support prevention and control efforts.

In November 2023, the National Health Commission issued the Healthy China Action-Cancer Prevention and Control Action Plan (2023-2030), which called for further improvement of the standardization, institutionalization, efficiency, and quality of cancer registration^2^. High-quality cancer registry data are essential for quantifying disease burden, identifying high-risk populations, and informing evidence-based prevention and control strategies.

Jiangsu Province is one of the earliest regions in China to establish population-based cancer registration, beginning in Qidong in 1972, and had achieved county- and district-wide coverage of follow-up cancer registration by the end of 2016^3^. Using population-based cancer registration data from Jiangsu Province from 2010 to 2021, this study aimed to assess the burden and temporal trends of five major gastrointestinal cancers, including esophageal cancer, gastric cancer, colon cancer, rectal cancer, and liver cancer. The study further compared patterns by sex and urban-rural residence, and projected their incidence and mortality to 2030.

## 2. Methods

### 2.1 Data source

This study used population-based cancer registration data from Jiangsu Province, China, from 2010 to 2021. The data were derived from the annual Jiangsu cancer report^4-13^. Only data from cancer registry sites in Jiangsu that met the quality requirements were used. The number of qualified registry sites increased from 27 in 2010 to 74 in 2021, including 33 urban registry sites and 41 rural registry sites in 2021 **(Supplement Table 1)**.

Cancer registration in Jiangsu is a systematic process for collecting, checking, storing, organizing, and statistically analyzing information on cancer incidence, death, and survival. Information on newly diagnosed cancer cases was mainly reported by medical institutions with cancer diagnosis and treatment capacity. Registry offices reviewed the submitted records, checked completeness and validity, and supplemented missing cases through follow-up and record linkage^13^.

Death information was obtained from death surveillance data and death certificates. Registry offices regularly checked deaths mentioning cancer as the underlying cause, contributing cause, or relevant disease history, and linked these records with the cancer registry database. Medical insurance records were also used as an auxiliary information source to identify cancer-related diagnoses and treatment records. Population denominators were obtained from public security and statistical departments^13^.

Incident cases and deaths were coded according to the International Classification of Diseases, 10th Revision (ICD-10). This study focused on five major gastrointestinal cancers: esophageal cancer (C15), gastric cancer (C16), colon cancer (C18), rectal cancer (C19-C20), and liver cancer (C22). The main outcomes were incidence and mortality. Annual incidence rates, mortality rates, age-specific rates, and age-standardized rates were extracted for each cancer type. Rates were expressed per 100,000 population. Age-standardized rates were calculated using the age distribution of the 2000 Chinese standard population based on the Fifth National Population Census^13^.

### 2.2 Quality control

Quality control was conducted throughout the cancer registration process. With reference to the data-quality evaluation framework by the National Cancer Center of China, the International Agency for Research on Cancer (IARC), and the International Association of Cancer Registries (IACR), submitted registry data were evaluated for completeness, validity, and comparability^14-16^.

The primary quality-control indicators included the proportion of morphologically verified cases (MV%), the proportion of death-certificate-only cases (DCO%), and the mortality-to-incidence ratio (M/I). From 2010 to 2021, MV% ranged from 68% to 74%, DCO% ranged from 0.61% to 1.21%, and the M/I ratio ranged from 0.68 to 0.74. All indicators met the quality requirements **(Supplement Table 2)**.

### 2.3 Statistical analysis

Microsoft Excel 2021 was used for data organization. Analyses were conducted by cancer type, outcome, sex, and residence. Age-standardized incidence and mortality rates were used as the principal measures for trend analysis.

Joinpoint regression was used for trend analysis. Calendar year was used as the independent variable, and the age-standardized incidence or mortality rate was used as the dependent variable. The annual percentage change (APC), average annual percentage change (AAPC), corresponding 95% confidence intervals (CIs), and P values were estimated.

Because the original source data contained a combined reporting period for 2011-2013, annual rates for 2011, 2012, and 2013 were imputed using a shifted linear interpolation approach. Provisional annual values for 2011, 2012, and 2013 were first linearly interpolated between the observed 2010 and 2014 values. The three interpolated values were then shifted by a constant so that their mean equaled the original combined 2011-2013 rate. The imputed annual rates were subsequently used in the Joinpoint analysis. For population denominators, the combined 2011-2013 person-years were allocated equally across the three years.

Forecasts were made for 2030. After Joinpoint regression, the APC and 95% CI from the most recent selected trend segment were extracted. The observed age-standardized rate in 2021 was used as the baseline, and projections assumed continuation of the most recent trend. The projected rate in year t was calculated as:

Rate_t = Rate_2021 x (1 + APC / 100)^^^(t - 2021)

The lower and upper projection bounds were calculated using the lower and upper limits of the APC 95% CI, respectively.

A two-sided *P* value of less than 0.05 was considered statistically significant. Statistical analyses were performed using Joinpoint Regression Program 6.0.1^17^ and Python 3.13.3.

## 3. Results

### 3.1 Trends in the surveyed population, 2010-2021

With the increasing number of qualified cancer registration sites, the surveyed population in Jiangsu increased substantially from 28,694,775 in 2010 to 74,669,580 in 2021 **(Supplement Figure 1A)**.

The male and female populations were generally balanced throughout the study period, with slightly fewer females than males. The surveyed populations in both urban and rural areas also increased over time, from 13,571,485 and 15,123,290 in 2010 to 33,125,493 and 41,544,087 in 2021, respectively **(Supplement Figure 1B, 1C)**.

### 3.2 Incidence and mortality of five major gastrointestinal cancers in 2021

In 2021, the overall age-standardized incidence rates of esophageal, gastric, colon, rectal, and liver cancer were 12.50, 17.81, 10.24, 7.98, and 11.77 per 100,000, respectively. For all five cancers, incidence rates were higher in males than in females, ranging from 1.46 to 2.83 times the corresponding female rates. By residence, urban residents had higher incidence rates for gastric, colon, and rectal cancer, whereas rural residents had higher rates for esophageal and liver cancer **(Table 1)**.

**Table 1.** Age-standardized incidence trends of five major gastrointestinal cancers in Jiangsu Province, 2010-2021.

| Stratum | Cancer | Age-standardized Incidence<br>Rate in 2010 (per 100,000) | Age-standardized Incidence<br>Rate in 2021 (per 100,000) | AAPC (95% CI) | P value |
| --- | --- | --- | --- | --- | --- |
| Overall | Esophageal cancer | 20.60 | 12.50 | -4.50 (-5.11, -3.29) | <0.001 |
|  | Gastric cancer | 22.21 | 17.81 | -2.13 (-3.41, -0.05) | 0.045 |
|  | Colon cancer | 4.20 | 10.24 | 8.13 (6.93, 10.23) | <0.001 |
|  | Rectal cancer | 5.08 | 7.98 | 3.72 (2.87, 5.42) | <0.001 |
|  | Liver cancer | 16.26 | 11.77 | -2.95 (-3.55, -1.86) | <0.001 |
| Male | Esophageal cancer | 28.72 | 18.43 | -4.05 (-4.62, -2.96) | <0.001 |
|  | Gastric cancer | 32.20 | 25.45 | -2.34 (-3.61, -0.57) | 0.006 |
|  | Colon cancer | 4.66 | 12.19 | 8.68 (6.89, 11.81) | <0.001 |
|  | Rectal cancer | 5.89 | 10.08 | 4.50 (3.41, 6.84) | <0.001 |
|  | Liver cancer | 24.51 | 17.48 | -3.12 (-3.82, -1.79) | <0.001 |
| Female | Esophageal cancer | 12.71 | 6.82 | -5.82 (-7.01, -3.53) | <0.001 |
|  | Gastric cancer | 12.64 | 10.58 | -1.63 (-2.76, -0.11) | 0.033 |
|  | Colon cancer | 3.76 | 8.36 | 7.34 (5.75, 10.42) | <0.001 |
|  | Rectal cancer | 4.32 | 5.96 | 2.49 (1.66, 3.99) | <0.001 |
|  | Liver cancer | 8.00 | 6.18 | -2.31 (-3.05, -0.83) | <0.001 |
| Urban | Esophageal cancer | 18.40 | 11.19 | -4.33 (-4.84, -3.23) | <0.001 |
|  | Gastric cancer | 26.17 | 18.83 | -3.13 (-3.92, -1.94) | <0.001 |
|  | Colon cancer | 6.01 | 12.18 | 6.13 (4.30, 9.24) | <0.001 |
|  | Rectal cancer | 5.89 | 8.56 | 3.12 (2.22, 5.35) | <0.001 |
|  | Liver cancer | 14.06 | 10.82 | -2.27 (-3.65, -0.31) | 0.024 |
| Rural | Esophageal cancer | 22.23 | 13.54 | -4.52 (-5.20, -3.35) | <0.001 |
|  | Gastric cancer | 19.25 | 16.99 | -1.22 (-3.68, 2.19) | 0.225 |
|  | Colon cancer | 2.86 | 8.69 | 9.63 (7.83, 15.10) | <0.001 |
|  | Rectal cancer | 4.47 | 7.52 | 4.59 (3.39, 7.03) | <0.001 |
|  | Liver cancer | 18.12 | 12.55 | -3.49 (-4.09, -2.44) | <0.001 |

In 2021, the overall age-standardized mortality rates of esophageal, gastric, colon, rectal, and liver cancer were 9.95, 12.45, 3.72, 3.56, and 10.65 per 100,000, respectively. Mortality rates were also higher in males than in females for all five cancers, ranging from 1.49 to 2.84 times the corresponding female rates. Urban residents had slightly higher mortality rates for gastric, colon, and rectal cancer, whereas rural residents had higher mortality rates for esophageal and liver cancer **(Table 2)**.

**Table 2.** Age-standardized mortality trends of five major gastrointestinal cancers in Jiangsu Province, 2010-2021.

| Stratum | Cancer | Age-standardized Mortality Rate<br>in 2010 (per 100,000) | Age-standardized Mortality<br>Rate in 2021 (per 100,000) | AAPC (95% CI) | P value |
| --- | --- | --- | --- | --- | --- |
| Overall | Esophageal cancer | 14.30 | 9.95 | -3.39 (-4.04, -2.45) | <0.001 |
|  | Gastric cancer | 15.34 | 12.45 | -2.08 (-3.04, -0.33) | 0.014 |
|  | Colon cancer | 1.68 | 3.72 | 7.21 (5.12, 10.33) | <0.001 |
|  | Rectal cancer | 2.76 | 3.56 | 2.16 (1.18, 3.87) | 0.002 |
|  | Liver cancer | 14.87 | 10.65 | -3.19 (-3.86, -2.05) | <0.001 |
| Male | Esophageal cancer | 19.87 | 14.88 | -2.80 (-3.54, -1.81) | <0.001 |
|  | Gastric cancer | 22.21 | 18.34 | -1.99 (-3.01, -0.25) | 0.020 |
|  | Colon cancer | 2.02 | 4.48 | 5.19 (4.41, 7.91) | <0.001 |
|  | Rectal cancer | 3.31 | 4.66 | 2.96 (1.62, 4.93) | <0.001 |
|  | Liver cancer | 22.28 | 15.86 | -3.30 (-4.10, -1.94) | <0.001 |
| Female | Esophageal cancer | 9.04 | 5.34 | -4.75 (-5.38, -3.86) | <0.001 |
|  | Gastric cancer | 8.96 | 7.04 | -2.26 (-4.09, 0.55) | 0.096 |
|  | Colon cancer | 1.38 | 3.01 | 7.04 (5.45, 10.70) | <0.001 |
|  | Rectal cancer | 2.28 | 2.53 | 0.73 (0.02, 1.73) | 0.044 |
|  | Liver cancer | 7.49 | 5.59 | -2.74 (-3.31, -1.67) | <0.001 |
| Urban | Esophageal cancer | 13.06 | 8.91 | -4.01 (-4.54, -3.32) | <0.001 |
|  | Gastric cancer | 18.09 | 12.58 | -3.55 (-4.27, -2.43) | <0.001 |
|  | Colon cancer | 2.25 | 4.51 | 5.70 (4.62, 8.04) | <0.001 |
|  | Rectal cancer | 3.12 | 3.73 | 1.38 (0.01, 3.33) | 0.050 |
|  | Liver cancer | 13.08 | 10.00 | -2.41 (-3.21, -0.88) | <0.001 |
| Rural | Esophageal cancer | 15.20 | 10.79 | -3.34 (-4.36, -1.82) | <0.001 |
|  | Gastric cancer | 13.37 | 12.35 | -1.02 (-3.13, 2.06) | 0.297 |
|  | Colon cancer | 1.27 | 3.08 | 6.39 (5.01, 11.13) | <0.001 |
|  | Rectal cancer | 2.48 | 3.42 | 2.88 (1.98, 4.86) | <0.001 |
|  | Liver cancer | 16.43 | 11.19 | -3.82 (-4.63, -2.82) | <0.001 |

### 3.3 Age-specific distribution of five major gastrointestinal cancers in 2021

In 2021, age-specific incidence rates for the five major gastrointestinal cancers increased markedly after 50 years of age, generally peaked at 80-84 years, and then declined slightly. Mortality showed a similar pattern for esophageal and gastric cancer, whereas mortality from colon, rectal, and liver cancer continued to rise into the oldest age groups **(Figure 1A, 1B)**.

**Figure 1.**
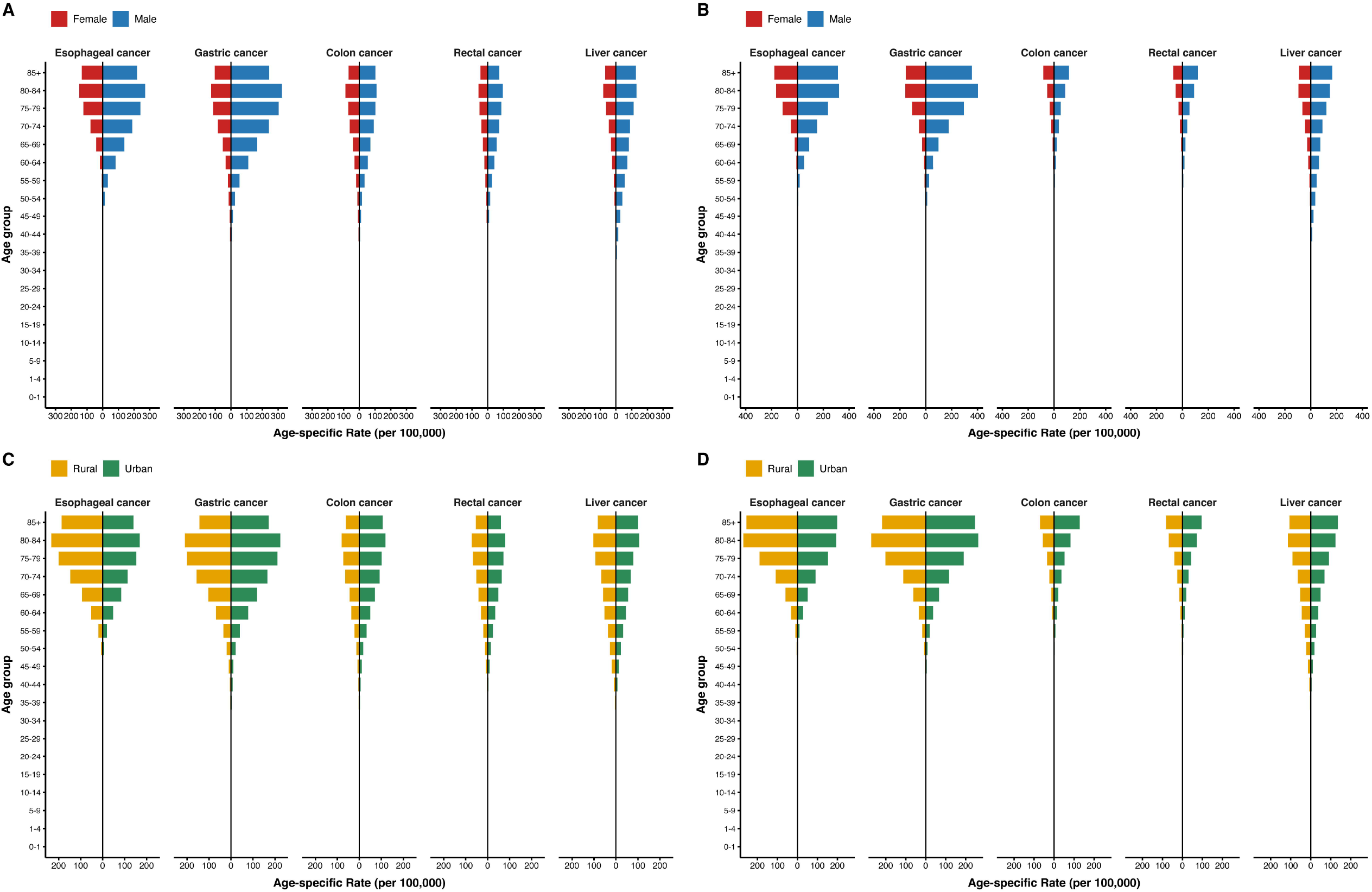
Age-specific incidence and mortality pyramids by sex and residence for five major gastrointestinal cancers in Jiangsu Province, 2021. (A) Incidence by sex; (B) mortality by sex; (C) incidence by residence; and (D) mortality by residence. Female and rural rates are shown on the left, and male and urban rates on the right.

Across nearly all age groups, males had higher incidence and mortality rates than females. Urban-rural differences varied by cancer type: urban populations generally had higher age-specific rates for gastric, colon, and rectal cancer, whereas rural populations tended to have higher rates for esophageal cancer **(Figure 1C, 1D)**.

### 3.4 Temporal trends in incidence and mortality of five major gastrointestinal cancers, 2010-2021

From 2010 to 2021, the overall age-standardized incidence rates of esophageal, gastric, and liver cancer declined in Jiangsu, with AAPCs of -4.50% (95% CI: -5.11%, -3.29%), -2.13% (95% CI: -3.41%, -0.05%), and -2.95% (95% CI: -3.55%, -1.86%), respectively. In contrast, the age-standardized incidence rates of colon and rectal cancer increased, with AAPCs of 8.13% (95% CI: 6.93%, 10.23%) and 3.72% (95% CI: 2.87%, 5.42%), respectively **(Table 1; Figure 2A)**.

**Figure 2.**
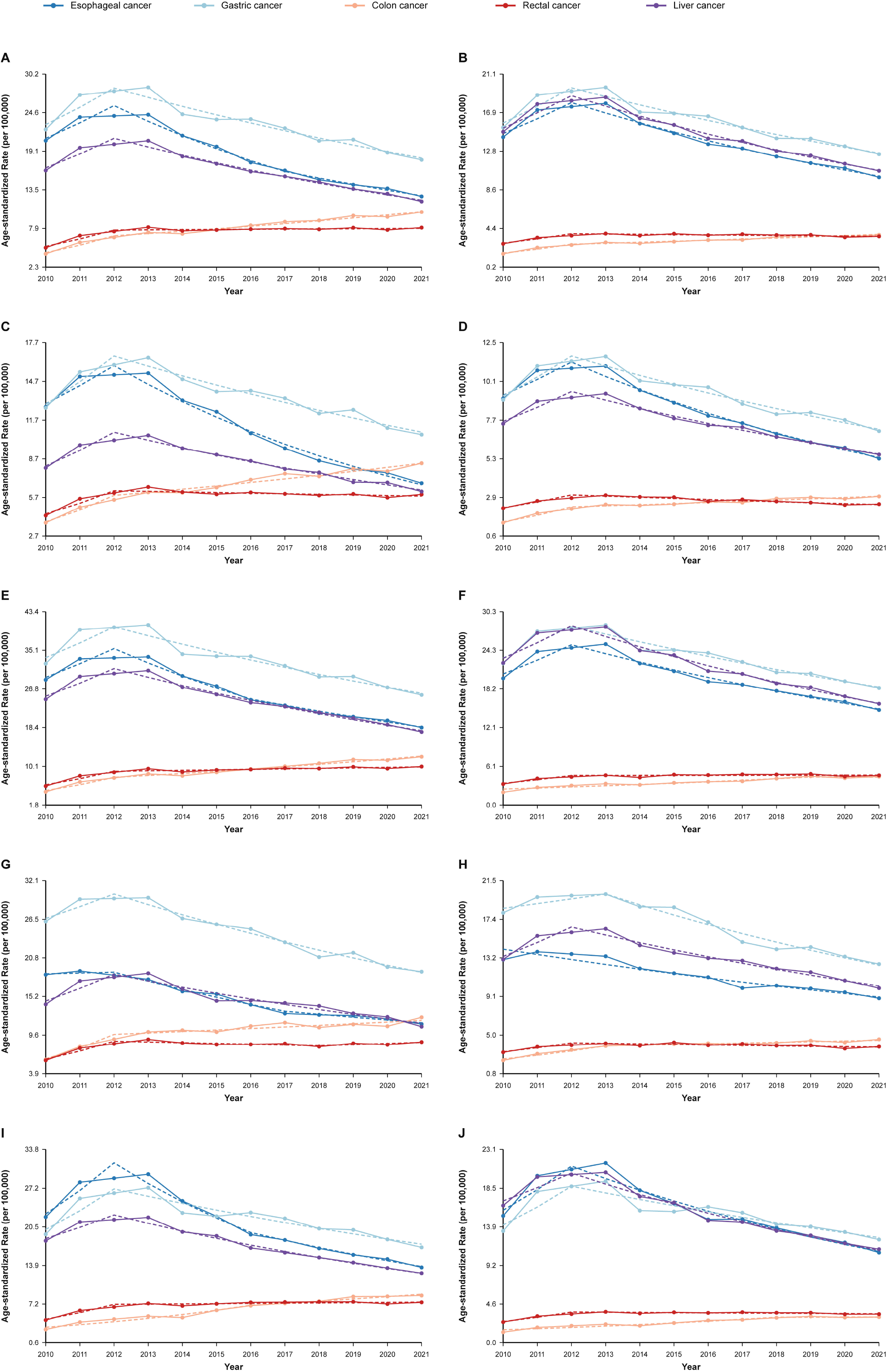
Joinpoint trends in incidence and mortality of five major gastrointestinal cancers across population subgroups in Jiangsu Province, 2010-2021. (A) Overall incidence; (B) overall mortality; (C) female incidence; (D) female mortality; (E) male incidence; (F) male mortality; (G) urban incidence; (H) urban mortality; (I) rural incidence; and (J) rural mortality. Solid lines indicate observed age-standardized rates and dashed lines indicate Joinpoint-fitted trends.

Segment-specific APC estimates showed an initial increase followed by a decline for esophageal, gastric, and liver cancer incidence. Colon cancer incidence increased throughout the study period, whereas rectal cancer incidence increased before leveling off after 2012 **(Supplement Table 3)**.

From 2010 to 2021, the overall age-standardized mortality rates of esophageal, gastric, and liver cancer decreased, with AAPCs of -3.39% (95% CI: -4.04%, -2.45%), -2.08% (95% CI: -3.04%, -0.33%), and -3.19% (95% CI: -3.86%, -2.05%), respectively. By contrast, mortality from colon and rectal cancer increased, with AAPCs of 7.21% (95% CI: 5.12%, 10.33%) and 2.16% (95% CI: 1.18%, 3.87%), respectively **(Table 2; Figure 2B)**. Segment-specific APC trends for mortality were broadly similar to those for incidence **(Supplement Table 4)**.

Trends were generally similar between males and females. For incidence, declines in gastric and liver cancer were steeper in males, whereas the decline in esophageal cancer was steeper in females; increases in colon and rectal cancer were greater in males. For mortality, the decline in esophageal cancer was steeper in females, whereas the decline in liver cancer was steeper in males. The increase in colon cancer mortality was greater in females, whereas the increase in rectal cancer mortality was greater in males **(Tables 1 and 2; Figure 2C-2F)**.

Residence-specific trends also differed by cancer type. For incidence, the decline in gastric cancer was steeper in urban areas, whereas the decline in liver cancer was steeper in rural areas; increases in colon and rectal cancer were greater in rural areas. For mortality, declines in esophageal and gastric cancer were steeper in urban areas, whereas the decline in liver cancer was steeper in rural areas. Increases in colon and rectal cancer mortality were greater in rural areas **(Tables 1 and 2; Figure 2G-2J)**.

### 3.5 Projected incidence and mortality of five major gastrointestinal cancers in 2030

Assuming continuation of the most recent Joinpoint segment, by 2030 the overall age-standardized incidence rates of esophageal, gastric, and liver cancer are projected to decline to 7.19, 11.41, and 6.73 per 100,000, respectively. Colon cancer incidence is projected to increase to 15.49 per 100,000, whereas rectal cancer incidence is projected to remain relatively stable at 8.24 per 100,000 **(Figure 3A)**.

**Figure 3.**
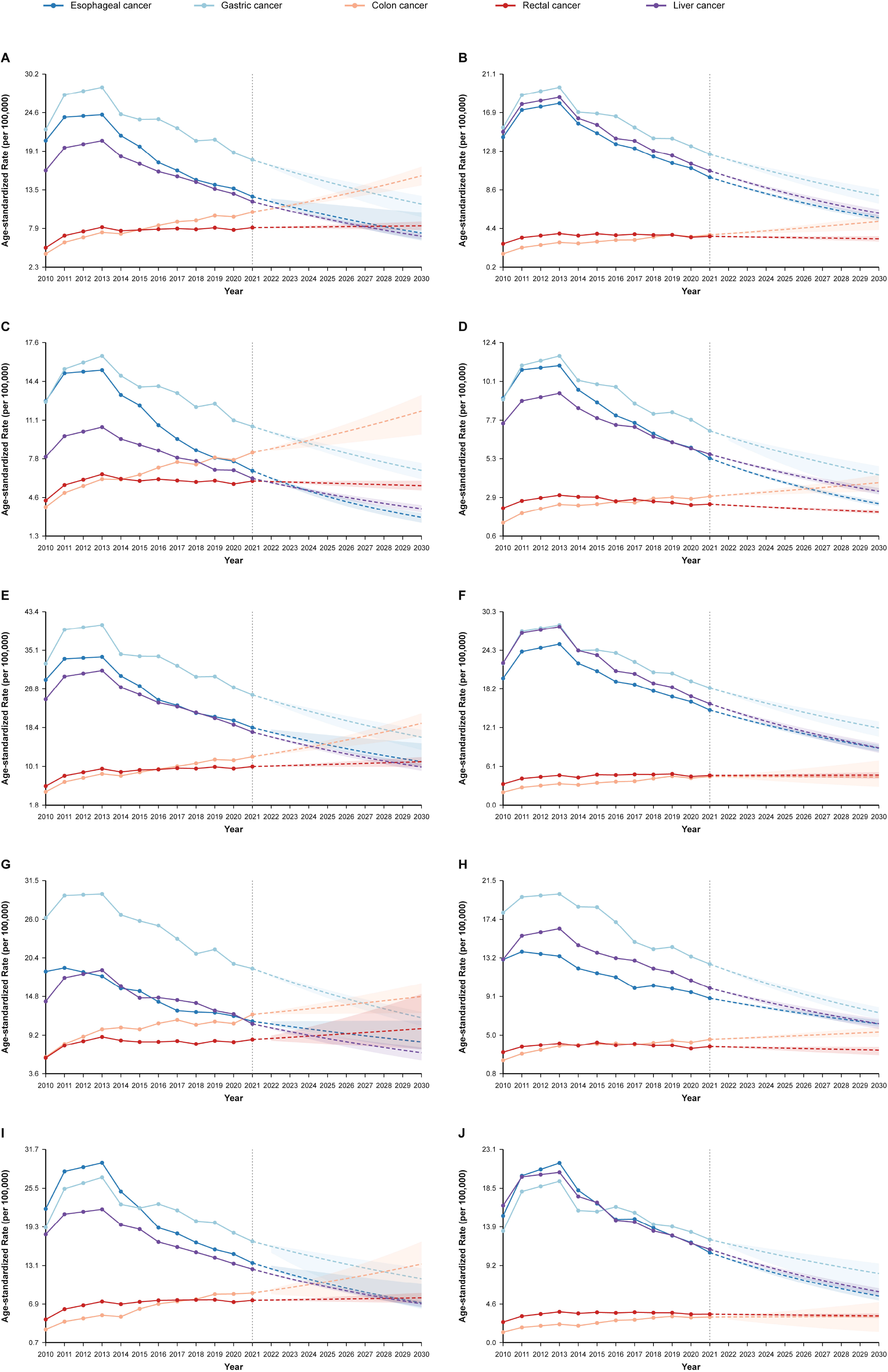
Forecasts of incidence and mortality of five major gastrointestinal cancers across population subgroups in Jiangsu Province. (A) Overall incidence; (B) overall mortality; (C) female incidence; (D) female mortality; (E) male incidence; (F) male mortality; (G) urban incidence; (H) urban mortality; (I) rural incidence; and (J) rural mortality. Solid lines indicate observed age-standardized rates through 2021, dashed lines indicate projected trends, and shaded bands indicate 95% confidence intervals.

By 2030, the overall age-standardized mortality rates of esophageal, gastric, and liver cancer are projected to decline to 5.55, 7.97, and 6.06 per 100,000, respectively. Colon cancer mortality is projected to increase to 5.17 per 100,000, whereas rectal cancer mortality is projected to remain relatively stable at 3.30 per 100,000 **(Figure 3B)**.

Projected patterns were generally similar between males and females, with lower incidence and mortality rates in females throughout 2030. Notably, colon cancer mortality is projected to increase in females but remain approximately stable in males. Urban and rural areas showed generally similar patterns overall **(Figure 3C-3J)**.

## 4. Discussion

Population-based cancer registry data provide an essential foundation for understanding the distribution and evolution of cancer burden across time, place, and population subgroups, and for translating epidemiologic evidence into prevention and control strategies. Using cancer registry data from Jiangsu Province, this study described the incidence and mortality patterns of five major digestive system cancers from 2010 to 2021, compared trends by sex and residence, and projected the burden to 2030. Overall, the findings suggest that Jiangsu is undergoing a clear transition in digestive cancer burden: esophageal cancer, gastric cancer, and liver cancer are declining, whereas colon cancer and, to a lesser extent, rectal cancer are increasing.

The first notable finding is the sustained decline in age-standardized incidence and mortality for esophageal, gastric, and liver cancers. In the overall population, the AAPCs for incidence were -4.50% for esophageal cancer, -2.13% for gastric cancer, and -2.95% for liver cancer, while the corresponding AAPCs for mortality were -3.39%, -2.08%, and -3.19%, respectively. These patterns are broadly consistent with national data showing long-term declines in the burden of upper gastrointestinal cancers and liver cancer in China^18-21^. This decline may be related to reduced exposure to traditional risk factors, improvements in living conditions and food storage, hepatitis B vaccination and liver disease control, and the gradual expansion of screening and early diagnosis in high-risk populations^20-23^. However, despite these favorable trends, gastric cancer remained the most prominent cancer in Jiangsu in 2021, with the highest age-standardized incidence (17.81/100,000) and mortality (12.45/100,000) among the five cancers examined. This suggests that Jiangsu still retains a substantial residual burden of upper gastrointestinal cancer, even as the overall pattern shifts.

In contrast, colon cancer emerged as a growing future burden. In the overall population, colon cancer showed the steepest increase in both incidence and mortality, with AAPCs of 8.13% and 7.21%, respectively, and the projected age-standardized incidence and mortality are expected to rise further to 15.50/100,000 and 5.17/100,000 by 2030. Rectal cancer also increased during 2010-2021, although the magnitude of change was smaller and the 2030 projection appeared comparatively stable. This pattern is consistent with the broader transition reported in China, where colorectal cancer has become a growing component of digestive cancer burden as the incidence of esophageal, gastric, and liver cancers declines ^18,24,25^. It is also compatible with local evidence from Wuxi (a city in Jiangsu) showing a substantial colorectal cancer mortality burden, suggesting that the growing colorectal burden is already relevant within Jiangsu^26^. The rising burden of colon and rectal cancer is likely associated with population aging, westernized dietary patterns, higher consumption of red and processed meat, obesity, physical inactivity, and other metabolic risk factors ^24,25,27^. The stronger and more sustained rise in colon cancer than in rectal cancer in our study suggests that colorectal prevention strategies in Jiangsu should not focus only on colorectal cancer as a combined entity, but should pay particular attention to the growing burden of colon cancer.

Another important finding is the marked age and sex disparity in burden. Across all five cancers, incidence and mortality increased substantially after the age of 50, and the burden was concentrated in older adults. This pattern is consistent with the biology of digestive cancers, which often result from long-term cumulative exposure to carcinogenic factors and the progressive accumulation of molecular damage over the life course^28,29^. It is also consistent with previous Chinese studies showing that colorectal and upper gastrointestinal cancer burdens are concentrated in middle-aged and older populations^18,24^. In addition, males consistently had higher incidence and mortality than females for all five cancers. The sex gap was especially apparent for esophageal, gastric, and liver cancers, but was also present for colon and rectal cancers. This male excess may be related to higher exposure to smoking and alcohol consumption, more frequent occupational and behavioral risk exposures, and a greater prevalence of chronic liver disease and other metabolic risk factors among men^18,27,30,31^. National analyses have similarly shown that smoking and alcohol remain major contributors to digestive cancer-related mortality in Chinese men^18^. These findings indicate that older adults, especially men aged 50 years and above, should continue to be priority populations for digestive cancer prevention, risk-factor modification, and screening.

The urban-rural comparison also deserves attention. Our results suggest that the growth rate of colon and rectal cancer burden was higher in rural than in urban residents. For incidence, the AAPC for colon cancer was 9.63% in rural areas versus 6.13% in urban areas, and for rectal cancer it was 4.59% versus 3.12%; for mortality, the corresponding AAPCs were 6.39% versus 5.70% for colon cancer and 2.88% versus 1.38% for rectal cancer. Although the absolute incidence and mortality rates of colon and rectal cancer were still higher in urban residents in 2021, the findings indicate that rural areas are catching up more rapidly, which may gradually narrow or even reverse historical urban-rural differences. This interpretation is supported by previous studies from China and local analyses from Jiangsu, which show substantial urban-rural disparities in cancer incidence^26,32,33^. Possible explanations for this pattern include a faster shift toward high-fat and low-fiber diets, increasing obesity and sedentary behavior, less equitable access to colonoscopy and early diagnosis, and weaker continuity of screening and treatment services in rural settings ^27,32,34^. Therefore, colorectal cancer prevention in Jiangsu should place special emphasis on strengthening rural screening capacity, referral pathways, and timely treatment.

The projection analysis further highlights the urgency of differentiated prevention strategies. If recent trends continue, the burden of esophageal cancer, gastric cancer, and liver cancer in Jiangsu is likely to keep declining through 2030, while colon cancer will continue to rise. These projections are compatible with previous Chinese forecasting studies, especially for liver cancer and colorectal cancer ^18,21,24,25^. From a public health perspective, this means Jiangsu faces a dual task: it must consolidate the gains already achieved in upper gastrointestinal and liver cancer control, while simultaneously addressing the growing challenge of colorectal cancer. In particular, the projected rise in colon cancer suggests that without stronger interventions on modifiable risk factors, the burden may continue to expand even if other digestive cancers decline.

This study has several strengths. First, it was based on real-world, population-based cancer registry data from Jiangsu Province, which provides a valuable picture of the province-level burden of digestive cancers. Second, the study compared patterns across sex and urban-rural residence groups, which helps identify high-risk populations and emerging inequalities. Third, the projection to 2030 offers forward-looking evidence to inform priority setting in cancer prevention and control.

Several limitations should also be acknowledged. First, the number of qualified cancer registries increased substantially from 2010 to 2021, and the covered population in 2021 was about 2.6 times that in 2010. Although this expansion could affect comparability over time, the main analyses were based on age-standardized rates, which should reduce, though not completely eliminate, this influence. Second, the original data for 2011-2013 were reported as a combined period, and annual values for those years were imputed for trend analysis. Although this approach allowed a continuous time-series analysis, some information loss and uncertainty are unavoidable. Third, the available data only covered 2010-2021, and more recent registry data have not yet been released. Longer observation periods and updated data will be needed to validate whether the projected trends persist in subsequent years.

In conclusion, this study indicates that the burden of digestive system cancers in Jiangsu Province is changing from a pattern dominated by esophageal, gastric, and liver cancers toward one in which colorectal cancer, especially colon cancer, is becoming increasingly prominent. While the continued decline in esophageal, gastric, and liver cancer is encouraging, gastric cancer still represented the highest burden among the five cancers in 2021, and the rapid increase in colon cancer deserves particular concern. Future prevention strategies in Jiangsu should prioritize adults aged 50 years and older, strengthen male-focused risk reduction, expand colorectal cancer screening and early diagnosis, improve rural prevention and treatment access, and continue consolidating existing gains in upper gastrointestinal and liver cancer control. Future studies should update these findings when newer registry data become available and further evaluate the effectiveness of targeted interventions in different population subgroups.

## Author Credit

Wei Wang: Data curation, Formal Analysis, Investigation, Methodology, Resources, Visualization, Writing – review & editing. Li Pan: Investigation, Writing – review & editing. Luqiu Tao: Data curation, Investigation, Writing – review & editing. Wei Wang: Conceptualization, Funding acquisition, Project administration, Writing – review & editing. Haiyan Ju: Investigation, Writing – review & editing. Hongru Zhu: Investigation, Writing – review & editing. Yanzheng Zou: Conceptualization, Data curation, Formal Analysis, Funding acquisition, Investigation, Software, Visualization, Writing – original draft.

## Ethics statement

This study used only publicly available, aggregate data extracted from previously published books and reports. No individual-level or identifiable human data were involved. Therefore, ethics committee or institutional review board approval was not required.

## Data availability statement

The data analyzed in this study were extracted from publicly available, previously published books and reports. All data were aggregate data and contained no individual-level or identifiable information. The original data sources are cited in the manuscript.

