## Supplementary Material for "Trends and Future Burden of Major Gastrointestinal Cancers in Jiangsu Province, China, 2010–2030"

| **Year** | **Overall** | **Urban** | **Rural** |
| --- | --- | --- | --- |
| **2010** | 27 | 11 | 18 |
| **2011** | 34 | 10 | 24 |
| **2012** | 34 | 10 | 24 |
| **2013** | 34 | 10 | 24 |
| **2014** | 32 | 9 | 23 |
| **2015** | 35 | 10 | 25 |
| **2016** | 41 | 10 | 31 |
| **2017** | 43 | 11 | 32 |
| **2018** | 48 | 19 | 29 |
| **2019** | 54 | 23 | 31 |
| **2020** | 69 | 30 | 39 |
| **2021** | 74 | 33 | 41 |

**Supplement Table 1.** The number of qualified registry sites in Jiangsu from 2010 to 2021

**Supplement Table 2.** The primary quality control indicators of the Jiangsu cancer registration process from 2010 to 2021

| **Year** | **M/I** | **DCO%** | **MV%** |
| --- | --- | --- | --- |
| **2010** | 0.69 | 1.21% | 68% |
| **2011-2013** | 0.68 | 0.97% | 67.55% |
| **2014** | 0.67 | 0.82% | 68.20% |
| **2015** | 0.67 | 0.61% | 68.70% |
| **2016** | 0.66 | 0.71% | 68.70% |
| **2017** | 0.64 | 1.07% | 70.04% |
| **2018** | 0.63 | 0.92% | 69.79% |
| **2019** | 0.61 | 0.72% | 70.41% |
| **2020** | 0.6 | 0.93% | 71.44% |
| **2021** | 0.57 | 0.62% | 73.55% |

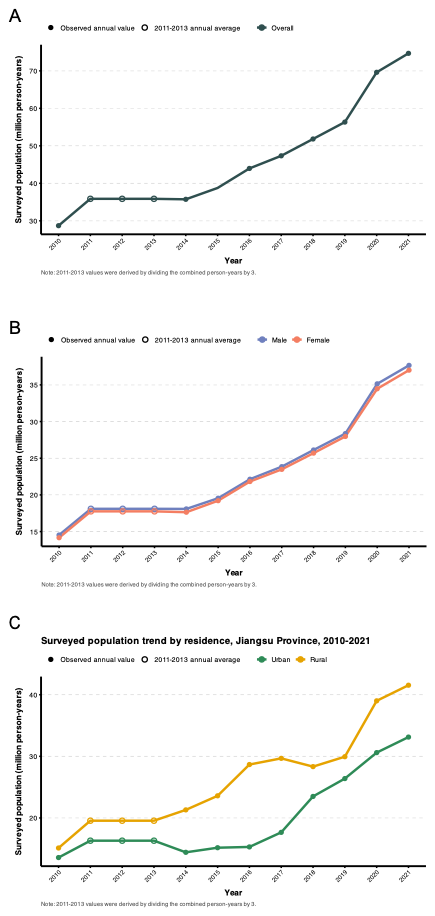

Supplement Figure 1. Surveyed population in Jiangsu from 2010 to 2021. (A) Overall; (B) By gender; (C) By residence.

**Supplement Table 3** Segment-specific APC estimates of incidence trends of five major gastrointestinal cancers from 2010 to 2021

| **Stratum** | **CancerName** | **Model** | **Segment** | **Segment Start** | **Segment End** | **APC** | **APC_LCL** | **APC_UCL** | **Significant** | **P Value** |
| --- | --- | --- | --- | --- | --- | --- | --- | --- | --- | --- |
| Overall | Esophageal cancer | 2 | 0 | 2010 | 2012 | 10.7663 | 2.1584 | 19.3556 | 1 | 0.017197 |
| Overall | Esophageal cancer | 2 | 1 | 2012 | 2017 | -8.8813 | -11.7391 | -7.4667 | 1 | 0 |
| Overall | Esophageal cancer | 2 | 2 | 2017 | 2021 | -5.9565 | -7.6188 | -2.2713 | 1 | 0.0012 |
| Overall | Gastric cancer | 1 | 0 | 2010 | 2012 | 10.9626 | -1.2049 | 25.3862 | 0 | 0.095981 |
| Overall | Gastric cancer | 1 | 1 | 2012 | 2021 | -4.8262 | -6.7585 | -3.7813 | 1 | 0.002 |
| Overall | Colon cancer | 1 | 0 | 2010 | 2012 | 24.9674 | 10.6138 | 39.4982 | 1 | 0 |
| Overall | Colon cancer | 1 | 1 | 2012 | 2021 | 4.7109 | 3.6306 | 5.6584 | 1 | 0.003599 |
| Overall | Rectal cancer | 1 | 0 | 2010 | 2012 | 20.3242 | 8.269 | 32.1278 | 1 | 0 |
| Overall | Rectal cancer | 1 | 1 | 2012 | 2021 | 0.3525 | -0.4968 | 1.1594 | 0 | 0.312338 |
| Overall | Liver cancer | 1 | 0 | 2010 | 2012 | 12.1537 | 4.0503 | 19.773 | 1 | 0.004799 |
| Overall | Liver cancer | 1 | 1 | 2012 | 2021 | -6.0215 | -6.7286 | -5.3772 | 1 | 0 |
| Male | Esophageal cancer | 2 | 0 | 2010 | 2012 | 10.1085 | 1.9921 | 17.8148 | 1 | 0.013597 |
| Male | Esophageal cancer | 2 | 1 | 2012 | 2016 | -8.7613 | -11.1095 | -6.9638 | 1 | 0 |
| Male | Esophageal cancer | 2 | 2 | 2016 | 2021 | -5.4562 | -6.5113 | -2.231 | 1 | 0.0008 |
| Male | Gastric cancer | 1 | 0 | 2010 | 2012 | 9.4875 | -1.839 | 21.5968 | 0 | 0.128374 |
| Male | Gastric cancer | 1 | 1 | 2012 | 2021 | -4.7902 | -6.6837 | -3.8164 | 1 | 0.0012 |
| Male | Colon cancer | 1 | 0 | 2010 | 2012 | 25.4626 | 8.7995 | 47.3798 | 1 | 0 |
| Male | Colon cancer | 1 | 1 | 2012 | 2021 | 5.269 | 3.4065 | 6.4951 | 1 | 0.019596 |
| Male | Rectal cancer | 1 | 0 | 2010 | 2012 | 21.2722 | 7.2156 | 37.941 | 1 | 0 |
| Male | Rectal cancer | 1 | 1 | 2012 | 2021 | 1.0948 | -0.0816 | 2.0607 | 0 | 0.065987 |
| Male | Liver cancer | 1 | 0 | 2010 | 2012 | 11.2178 | 2.1652 | 20.3957 | 1 | 0.015197 |
| Male | Liver cancer | 1 | 1 | 2012 | 2021 | -6.0451 | -6.92 | -5.316 | 1 | 0 |
| Female | Esophageal cancer | 1 | 0 | 2010 | 2012 | 10.7597 | -3.446 | 27.1102 | 0 | 0.171566 |
| Female | Esophageal cancer | 1 | 1 | 2012 | 2021 | -9.1489 | -10.8361 | -8.01 | 1 | 0 |
| Female | Gastric cancer | 1 | 0 | 2010 | 2012 | 13.5284 | 1.4447 | 24.2602 | 1 | 0.023195 |
| Female | Gastric cancer | 1 | 1 | 2012 | 2021 | -4.7158 | -5.9089 | -3.7962 | 1 | 0 |
| Female | Colon cancer | 1 | 0 | 2010 | 2012 | 23.9208 | 7.8315 | 45.033 | 1 | 0 |
| Female | Colon cancer | 1 | 1 | 2012 | 2021 | 3.9729 | 1.8935 | 5.2168 | 1 | 0.025995 |
| Female | Rectal cancer | 1 | 0 | 2010 | 2012 | 18.4536 | 7.2146 | 28.7178 | 1 | 0 |
| Female | Rectal cancer | 1 | 1 | 2012 | 2021 | -0.755 | -1.5673 | 0.0314 | 0 | 0.059988 |
| Female | Liver cancer | 1 | 0 | 2010 | 2012 | 14.9299 | 3.746 | 25.6769 | 1 | 0.004799 |
| Female | Liver cancer | 1 | 1 | 2012 | 2021 | -5.7778 | -6.7144 | -4.9201 | 1 | 0 |
| Urban | Esophageal cancer | 2 | 0 | 2010 | 2012 | 0.6613 | -4.1649 | 7.66 | 0 | 0.987403 |
| Urban | Esophageal cancer | 2 | 1 | 2012 | 2017 | -6.9896 | -10.1216 | -5.569 | 1 | 0 |
| Urban | Esophageal cancer | 2 | 2 | 2017 | 2021 | -3.378 | -4.9415 | 0.0988 | 0 | 0.054789 |
| Urban | Gastric cancer | 1 | 0 | 2010 | 2012 | 6.4939 | -1.7937 | 14.3172 | 0 | 0.161568 |
| Urban | Gastric cancer | 1 | 1 | 2012 | 2021 | -5.1497 | -6.2455 | -4.4251 | 1 | 0 |
| Urban | Colon cancer | 1 | 0 | 2010 | 2012 | 25.9945 | 7.1663 | 47.8746 | 1 | 0 |
| Urban | Colon cancer | 1 | 1 | 2012 | 2021 | 2.1654 | 0.1985 | 3.5554 | 1 | 0.045991 |
| Urban | Rectal cancer | 2 | 0 | 2010 | 2012 | 19.6607 | 7.5042 | 35.8336 | 1 | 0 |
| Urban | Rectal cancer | 2 | 1 | 2012 | 2018 | -1.2745 | -5.6022 | 0.2828 | 0 | 0.087582 |
| Urban | Rectal cancer | 2 | 2 | 2018 | 2021 | 1.8965 | -1.9961 | 6.5324 | 0 | 0.117177 |
| Urban | Liver cancer | 1 | 0 | 2010 | 2012 | 12.6698 | -0.6824 | 26.9314 | 0 | 0.071986 |
| Urban | Liver cancer | 1 | 1 | 2012 | 2021 | -5.3149 | -7.1921 | -4.1662 | 1 | 0.0008 |
| Rural | Esophageal cancer | 2 | 0 | 2010 | 2012 | 17.7028 | 5.9661 | 26.686 | 1 | 0.0004 |
| Rural | Esophageal cancer | 2 | 1 | 2012 | 2016 | -11.267 | -14.025 | -8.9932 | 1 | 0 |
| Rural | Esophageal cancer | 2 | 2 | 2016 | 2021 | -6.8855 | -8.1054 | -3.1563 | 1 | 0 |
| Rural | Gastric cancer | 1 | 0 | 2010 | 2012 | 16.1061 | -1.3541 | 41.1908 | 0 | 0.09878 |
| Rural | Gastric cancer | 1 | 1 | 2012 | 2021 | -4.7042 | -10.7648 | -3.1953 | 1 | 0.006799 |
| Rural | Colon cancer | 1 | 0 | 2010 | 2016 | 13.7405 | 10.408 | 50.1003 | 1 | 0 |
| Rural | Colon cancer | 1 | 1 | 2016 | 2021 | 4.9026 | -2.2572 | 7.6994 | 0 | 0.084783 |
| Rural | Rectal cancer | 1 | 0 | 2010 | 2012 | 24.7283 | 8.1545 | 42.3183 | 1 | 0 |
| Rural | Rectal cancer | 1 | 1 | 2012 | 2021 | 0.5744 | -0.6708 | 1.6369 | 0 | 0.29994 |
| Rural | Liver cancer | 1 | 0 | 2010 | 2012 | 10.4463 | 2.4112 | 17.8809 | 1 | 0.009998 |
| Rural | Liver cancer | 1 | 1 | 2012 | 2021 | -6.3385 | -7.0706 | -5.7115 | 1 | 0 |

**Supplement Table 4** Segment-specific APC estimates of mortality trends of five major gastrointestinal cancers from 2010 to 2021

| **Stratum** | **Cancer Name** | **Model** | **Segment** | **Segment Start** | **Segment End** | **APC** | **APC_LCL** | **APC_UCL** | **Significant** | **P Value** |
| --- | --- | --- | --- | --- | --- | --- | --- | --- | --- | --- |
| Overall | Esophageal cancer | 1 | 0 | 2010 | 2012 | 10.7961 | 2.6871 | 17.2165 | 1 | 0.005999 |
| Overall | Esophageal cancer | 1 | 1 | 2012 | 2021 | -6.2829 | -6.8809 | -5.7026 | 1 | 0 |
| Overall | Gastric cancer | 1 | 0 | 2010 | 2012 | 11.3906 | 0.1363 | 23.5324 | 1 | 0.045991 |
| Overall | Gastric cancer | 1 | 1 | 2012 | 2021 | -4.8394 | -6.0325 | -3.9519 | 1 | 0 |
| Overall | Colon cancer | 1 | 0 | 2010 | 2012 | 24.3795 | 6.9118 | 46.4761 | 1 | 0 |
| Overall | Colon cancer | 1 | 1 | 2012 | 2021 | 3.7321 | 1.491 | 5.0183 | 1 | 0.031994 |
| Overall | Rectal cancer | 1 | 0 | 2010 | 2012 | 16.852 | 4.6759 | 28.419 | 1 | 0 |
| Overall | Rectal cancer | 1 | 1 | 2012 | 2021 | -0.8477 | -1.8549 | 0.0159 | 0 | 0.054789 |
| Overall | Liver cancer | 1 | 0 | 2010 | 2012 | 10.9024 | 2.0918 | 18.8318 | 1 | 0.011598 |
| Overall | Liver cancer | 1 | 1 | 2012 | 2021 | -6.0677 | -6.8282 | -5.3897 | 1 | 0 |
| Male | Esophageal cancer | 1 | 0 | 2010 | 2012 | 10.5563 | 2.0261 | 17.3373 | 1 | 0.012398 |
| Male | Esophageal cancer | 1 | 1 | 2012 | 2021 | -5.5433 | -6.2003 | -4.9418 | 1 | 0 |
| Male | Gastric cancer | 1 | 0 | 2010 | 2012 | 10.3742 | -0.5873 | 22.2432 | 0 | 0.065187 |
| Male | Gastric cancer | 1 | 1 | 2012 | 2021 | -4.549 | -5.7981 | -3.6809 | 1 | 0.0008 |
| Male | Colon cancer | 1 | 0 | 2010 | 2019 | 6.4152 | 5.3995 | 19.8688 | 1 | 0.003199 |
| Male | Colon cancer | 1 | 1 | 2019 | 2021 | -0.1487 | -4.8165 | 5.0573 | 0 | 0.784643 |
| Male | Rectal cancer | 1 | 0 | 2010 | 2012 | 16.756 | 3.7393 | 30.1935 | 1 | 0.0004 |
| Male | Rectal cancer | 1 | 1 | 2012 | 2021 | 0.1274 | -1.304 | 1.1198 | 0 | 0.869426 |
| Male | Liver cancer | 1 | 0 | 2010 | 2012 | 10.6111 | 0.9749 | 20.1443 | 1 | 0.029994 |
| Male | Liver cancer | 1 | 1 | 2012 | 2021 | -6.1405 | -7.095 | -5.3552 | 1 | 0 |
| Female | Esophageal cancer | 1 | 0 | 2010 | 2012 | 10.5029 | 2.5324 | 16.818 | 1 | 0.015597 |
| Female | Esophageal cancer | 1 | 1 | 2012 | 2021 | -7.8474 | -8.4373 | -7.2813 | 1 | 0 |
| Female | Gastric cancer | 1 | 0 | 2010 | 2012 | 12.5429 | -1.9137 | 32.1757 | 0 | 0.122775 |
| Female | Gastric cancer | 1 | 1 | 2012 | 2021 | -5.2792 | -8.531 | -4.0442 | 1 | 0.003999 |
| Female | Colon cancer | 1 | 0 | 2010 | 2012 | 28.5349 | 8.8916 | 55.5481 | 1 | 0 |
| Female | Colon cancer | 1 | 1 | 2012 | 2021 | 2.7743 | 1.0978 | 4.0834 | 1 | 0.023595 |
| Female | Rectal cancer | 1 | 0 | 2010 | 2012 | 16.1072 | 5.2252 | 22.7676 | 1 | 0 |
| Female | Rectal cancer | 1 | 1 | 2012 | 2021 | -2.3981 | -3.0353 | -1.7536 | 1 | 0 |
| Female | Liver cancer | 1 | 0 | 2010 | 2012 | 11.4064 | 3.4496 | 18.8066 | 1 | 0.005199 |
| Female | Liver cancer | 1 | 1 | 2012 | 2021 | -5.6324 | -6.2603 | -5.0257 | 1 | 0 |
| Urban | Esophageal cancer | 0 | 0 | 2010 | 2021 | -4.0101 | -4.5363 | -3.3224 | 1 | 0 |
| Urban | Gastric cancer | 1 | 0 | 2010 | 2013 | 2.6764 | -1.9684 | 12.2242 | 0 | 0.333933 |
| Urban | Gastric cancer | 1 | 1 | 2013 | 2021 | -5.7817 | -6.8427 | -4.9784 | 1 | 0 |
| Urban | Colon cancer | 1 | 0 | 2010 | 2013 | 16.8131 | 8.8961 | 37.2201 | 1 | 0 |
| Urban | Colon cancer | 1 | 1 | 2013 | 2021 | 1.8163 | 0.6788 | 2.9097 | 1 | 0.011598 |
| Urban | Rectal cancer | 1 | 0 | 2010 | 2012 | 13.9958 | 1.7117 | 27.3439 | 1 | 0.006799 |
| Urban | Rectal cancer | 1 | 1 | 2012 | 2021 | -1.2321 | -3.1489 | -0.1812 | 1 | 0.021996 |
| Urban | Liver cancer | 1 | 0 | 2010 | 2012 | 11.1498 | 0.8128 | 21.8306 | 1 | 0.027994 |
| Urban | Liver cancer | 1 | 1 | 2012 | 2021 | -5.1969 | -6.1867 | -4.3606 | 1 | 0 |
| Rural | Esophageal cancer | 1 | 0 | 2010 | 2012 | 15.4282 | 2.3657 | 26.5544 | 1 | 0.013197 |
| Rural | Esophageal cancer | 1 | 1 | 2012 | 2021 | -7.0736 | -8.0887 | -6.1666 | 1 | 0 |
| Rural | Gastric cancer | 1 | 0 | 2010 | 2012 | 15.2989 | -0.9763 | 37.9153 | 0 | 0.088782 |
| Rural | Gastric cancer | 1 | 1 | 2012 | 2021 | -4.3245 | -7.8648 | -2.8862 | 1 | 0.006799 |
| Rural | Colon cancer | 1 | 0 | 2010 | 2018 | 8.8063 | 7.1791 | 31.5852 | 1 | 0.0008 |
| Rural | Colon cancer | 1 | 1 | 2018 | 2021 | 0.2142 | -9.0961 | 5.1916 | 0 | 0.85223 |
| Rural | Rectal cancer | 1 | 0 | 2010 | 2012 | 20.4069 | 7.1291 | 34.2507 | 1 | 0 |
| Rural | Rectal cancer | 1 | 1 | 2012 | 2021 | -0.6533 | -1.6306 | 0.229 | 0 | 0.141172 |
| Rural | Liver cancer | 1 | 0 | 2010 | 2012 | 9.3871 | 0.2776 | 16.2769 | 1 | 0.045191 |
| Rural | Liver cancer | 1 | 1 | 2012 | 2021 | -6.529 | -7.3734 | -5.8444 | 1 | 0 |
